# Complex Emotion Recognition in Neurological and Neurodevelopmental Conditions

**DOI:** 10.64898/2025.11.26.25341046

**Authors:** Rebecca Johannessen, Marcel Eicher, Anne Hansen, Michelle Regli, Lesley Ramseier, Tamara Hibbert, Hannah Sievers, Marah Ruepp, Sarah Scheibelhofer, Lukas Imbach, Hennric Jokeit

## Abstract

Social cognition deficits are prevalent but under-recognised in neurological and neurodevelopmental disorders. Complex emotion recognition (CER) – encompassing mentalising, theory of mind, and perspective-taking – is crucial for social functioning but lacks a standardised clinical assessment. We evaluated CER performance across neurological and neurodevelopmental populations using a novel digital screening instrument. A novel online assessment tool using muted video clips of dyadic interactions was administered to six clinical cohorts compared to controls (HC, N = 449): temporal lobe epilepsy (TLE, N = 36), multiple sclerosis (MS, N = 64), autism spectrum disorder (ASD, N = 90), attention deficit hyperactivity disorder (ADHD, N = 54), and healthy adults aged >65 years (H65+, N = 35). Reduced CER scores were observed across all clinical groups compared with HC (n = 455): ASD (Hedges’ *g* = −1.12, 95% CI −1.35 to −0.88), TLE (*g* = −0.93, 95% CI −1.29 to −0.60), H65+ (*g* = −0.73, 95% CI −1.07 to −0.38), MS (*g* = −0.65, 95% CI −0.91 to −0.38), and ADHD (*g* = −0.35, 95% CI −0.63 to −0.06). Significantly reduced scores were most prevalent for ASD (37%), H65+ (28%), and TLE (19%). Complex emotion recognition deficits are pervasive across clinical conditions, with the highest prevalence in individuals with ASD, TLE, and H65+. Our findings demonstrate the clinical importance of routine CER screening in these populations. The COSIMO test provides a practical solution for identifying patients at risk of social cognitive impairment who may benefit from counselling and targeted interventions.

## 1 INTRODUCTION

Social cognition—the ability to recognise and interpret social signals—depends on cortical networks involving the limbic, temporal, frontal, and prefrontal regions (Broicher et al., 2012; Fusar-Poli et al., 2009; Pitcher et al., 2023; Pitcher and Ungerleider, 2020; Wolf et al., 2010). Given the distributed nature of these networks, social cognitive impairments are observed in various psychiatric and neurological conditions (Cotter et al., 2018), contributing to reduced social functioning and quality of life (Marafioti et al., 2024; Steiger and Jokeit, 2017; Wang et al., 2015). Despite growing evidence that social cognition represents a transdiagnostic marker of neuropathological and psychopathological mechanisms (Cotter et al., 2018), it remains consistently underassessed in clinical diagnostics.

Direct cross-condition comparisons using methodologically consistent, valid paradigms are necessary to investigate the effects of different brain pathologies on social cognition. Basic emotion recognition tasks that employ static facial or prosodic stimuli primarily index temporal–limbic processes, as in temporal lobe epilepsy (TLE) (Bauer et al., 2023; Eicher and Jokeit, 2022; Qi et al., 2022; Wang et al., 2022; Ziaei et al., 2023), but lack ecological validity, i.e. relevance to everyday functioning. On the other hand, mentalising or theory of mind tasks are often used to assess higher-order social cognition in populations with prefrontal pathology, such as attention deficit hyperactivity disorder (ADHD) (Bora and Pantelis, 2016; Borhani and Nejati, 2018) or age-related cortical atrophy (Grainger et al., 2023; Henry et al., 2022; Roheger et al., 2022), as well as in conditions involving distributed network diversity or dysfunction, including autism spectrum disorder (ASD) (Bora and Pantelis, 2016; Cotter et al., 2018; Lievore et al., 2023; Yeung, 2022) or multiple sclerosis (MS) (Chalah and Ayache, 2017; Cotter et al., 2016; Lin et al., 2021; Pöttgen et al., 2013). In contrast to basic emotion tasks, higher-order social cognition tasks often boast a higher ecological validity, but also rely on multiple cognitive functions, including working memory and language comprehension, limiting their suitability for neurological populations with complex cognitive impairments (Eicher and Jokeit, 2022; Jokeit et al., 2018; Steiger and Jokeit, 2017). Consequently, heterogeneity in task design limits the comparability of findings across disorders (Cotter et al., 2018; Eicher and Jokeit, 2022).

Dynamic paradigms assessing complex emotion recognition (CER) bridge basic and higher-order social cognitive approaches by integrating dynamic facial expressions, body language, and social context to infer complex emotions, thereby capturing the multidimensional nature of real-world social interactions. In this study, we used a single video-based CER paradigm to compare the severity and prevalence of social cognitive impairments across TLE, MS, ASD, ADHD, and healthy agers aged 65 years and above (H65+) with healthy controls under 65 (HC). In doing so, we assessed the impact of different pathologies on complex emotion recognition and investigated its potential as a transdiagnostic marker.

## 2 METHODS

### 2.1 Participants

Assessments took place either online or in a clinical setting at the Swiss Epilepsy Centre. Clinically tested participants underwent routine neuropsychological assessment or were tested during their inpatient stay. We recruited participants through university mailing lists, elderly collectives, organisational forums, and personal contacts. Diagnoses relied on existing patient reports for clinically tested participants and self-reports for online participants. Anyone aged 16 years or older was welcomed to participate.

Online participants confirmed their consent to participate before completing the demographic survey and test. All in-person participants provided written informed consent. A 1 CHF donation was given to charity for each online participation, whereas in-person HC and H65+ participants were compensated for their travel costs with 15 CHF.

The data included in this study were collected between September 2022 and June 2025 with a stepwise launch for new clinical cohorts. We strived for 50-100 participants per group and 500 HC (for the purpose of collecting normative data). Data collection was based on a continuous testing approach, providing a growing pool of participants for future studies to draw from, as well as normative data for clinical practice. While we tested individuals with TLE in person, ASD and MS were recruited and tested online only, and H65+ and ADHD were mixed groups with both online and in-person participants.

### 2.2 Materials

#### 2.2.1 Complex Emotion Recognition Paradigm

Twenty-five muted videos of complex dyadic interactions (figure 1) were shown using a tablet by a clinician or completed independently online by the participants. Participants were shown one clip at a time with a yellow frame around the left or right half of the video, indicating the character on which they should focus. They were then asked to select the emotion that best fit the character in focus. The emotional words describing the characters were chosen to force a clear understanding of the short interactions and included words that describe the complex emotions of either an active (e.g. “joking”) or reactive (e.g. “amused”) nature. Two options had the correct valence and the third option had the opposite valence. Participants received one point if they could identify the most appropriate option. After the participant responded to one item, the next video was played. Each video could be repeated once. The test took 5-7 minutes to complete.

**Figure 1.**
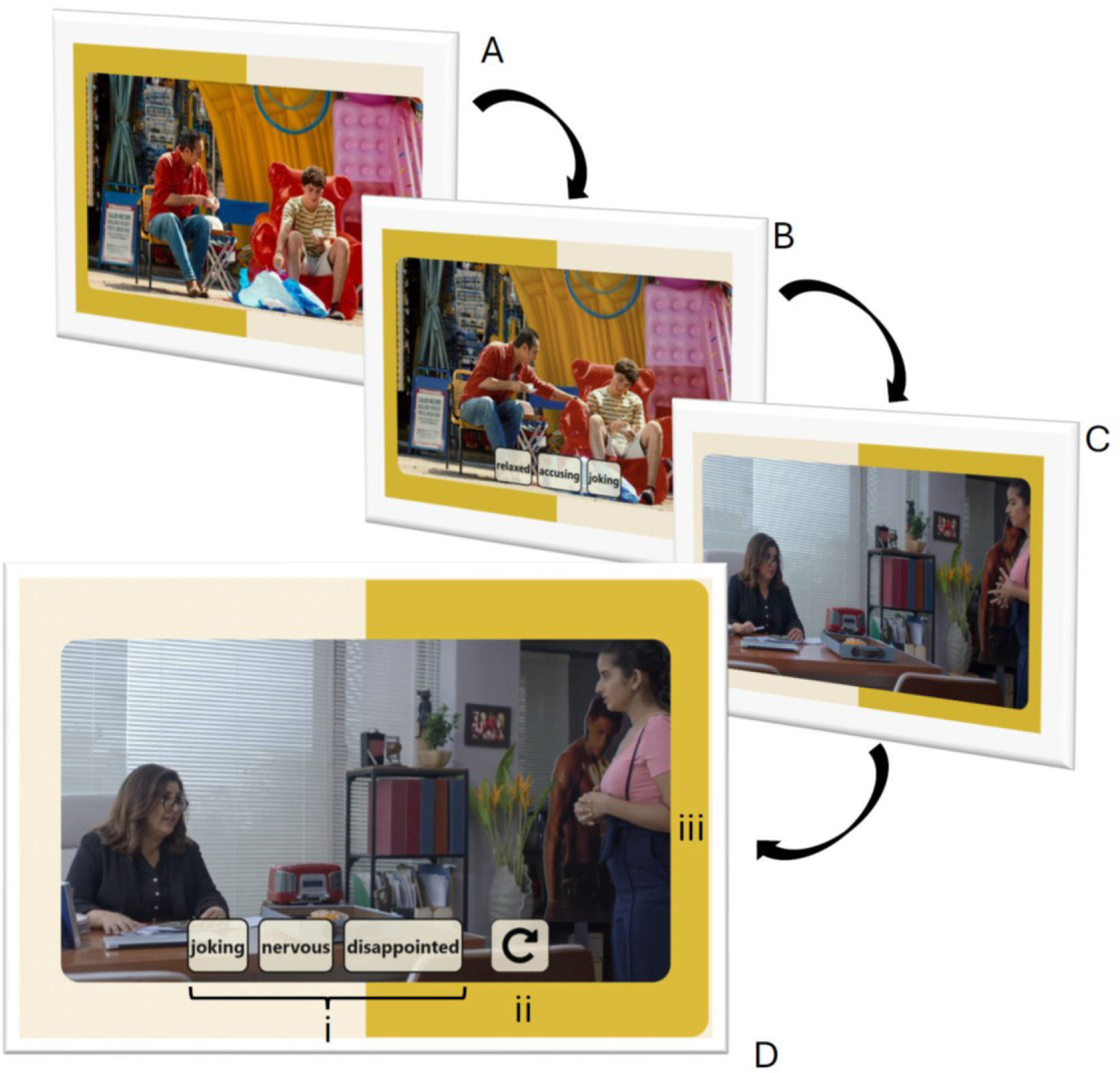
Example items from COSIMO. *(A) The first video was played automatically when the test started. (B) After the video, three options appear at the bottom of a still image of the final frame. (C) As soon as a participant responds, the next video plays automatically until all 25 items have been responded to. (D) Participants could select one of three options, either by touch or using a cursor (i). If they need to, they may watch the video again once using the repeat button (ii). A yellow frame (iii) indicates whether they should focus on the left- or right-hand character*.

#### 2.2.2 Development of the CER paradigm

The paradigm was developed from a collection of 600 muted, 4-8 second long video clips from streamed series and films, with a focus on the demographic diversity of the actors. The emotional words were adapted from the Reading the Mind in the Eyes Test (Baron-Cohen et al., 2001) and simplified by excluding less-used near synonyms during a pilot study. The items were developed using responses from 582 participants from Amazon Mechanical Turk in English. A list of 75 videos was then reviewed and translated into German in a panel study. Two versions with 25 videos were released in September 2022 as pilot versions for interested practitioners and researchers in the form of an independent web application named COSIMO (https://cosimo-project.com).

This study only reports on one of the two COSIMO test versions, as a comparison of test versions is beyond the scope of this paper. Based on odds ratio comparisons, we eliminated six items from the final analyses because of anomalous response patterns, where TLE or ASD more frequently responded with the “correct” answer than HC. An overview of the psychometric validation and norming of the German version of COSIMO is in development and will be published elsewhere (Johannessen et al., n.d.).

#### 2.2.3 Additional tests and surveys

In-person participants completed the Montreal Cognitive Assessment (MoCA) (Nasreddine et al., 2005), a well-validated screening tool for general cognitive disability, or were assessed using a more extensive neuropsychological test battery. Because of the heterogeneity of the other tests used, we chose to use MoCA to investigate the effect of general cognitive disability in the subset of patients who had completed this task.

Online participants completed a demographic survey, from which age, education, gender, and diagnosis were extracted for the purposes of this study. The same information was collected orally from in-person participants or extracted from patient reports.

### 2.3 Data Analysis

We extracted all German-speaking responses from online participants or patients at the Swiss Epilepsy Centre from the COSIMO database before importing them into R version 4.5.1. We included each instance in one of the study groups based on the recruitment source and diagnostic information or excluded them if the age or diagnosis was missing or if the diagnosis did not correspond to any of the clinical groups in this study.

In preparation of parametric testing, we checked for normal distribution in age, COSIMO score, and MoCA scores using the Shapiro-Wilk test and visual inspection of the density plots, and transformed the variables using Tukey Ladder Transformation if deemed necessary.(Mangiafico, 2015)

For the main analysis, effect sizes between all groups and healthy controls (HC) were calculated using Hedge’s *g* and their significance at a 95% confidence level using Welch’s two-sample t-test. We chose a more conservative two-tailed t-test because the paradigm used was novel. Additionally, we calculated the prevalence of having a result below −1.5 SD or −2 SD for each group.

## 3 RESULTS

In total, 890 participants completed COSIMO in German. We assigned 728 participants to one of the groups. The final sample included 449 healthy controls below 65 (HC), 35 healthy agers 65 and above (H65+), 64 participants with Multiple Sclerosis (MS), 35 participants with Temporal Lobe Epilepsy (TLE), 90 participants with Autism Spectrum Disorder (ASD), and 54 participants with Attention Deficit and Hyperactivity Disorder (ADHD). Additional demographic information is presented in Table 1. The remaining participants (N = 145) were excluded because they had a diagnosis that was not included in the study (N = 143) or had unlikely random response patterns (N = 2).

**Table 1.**
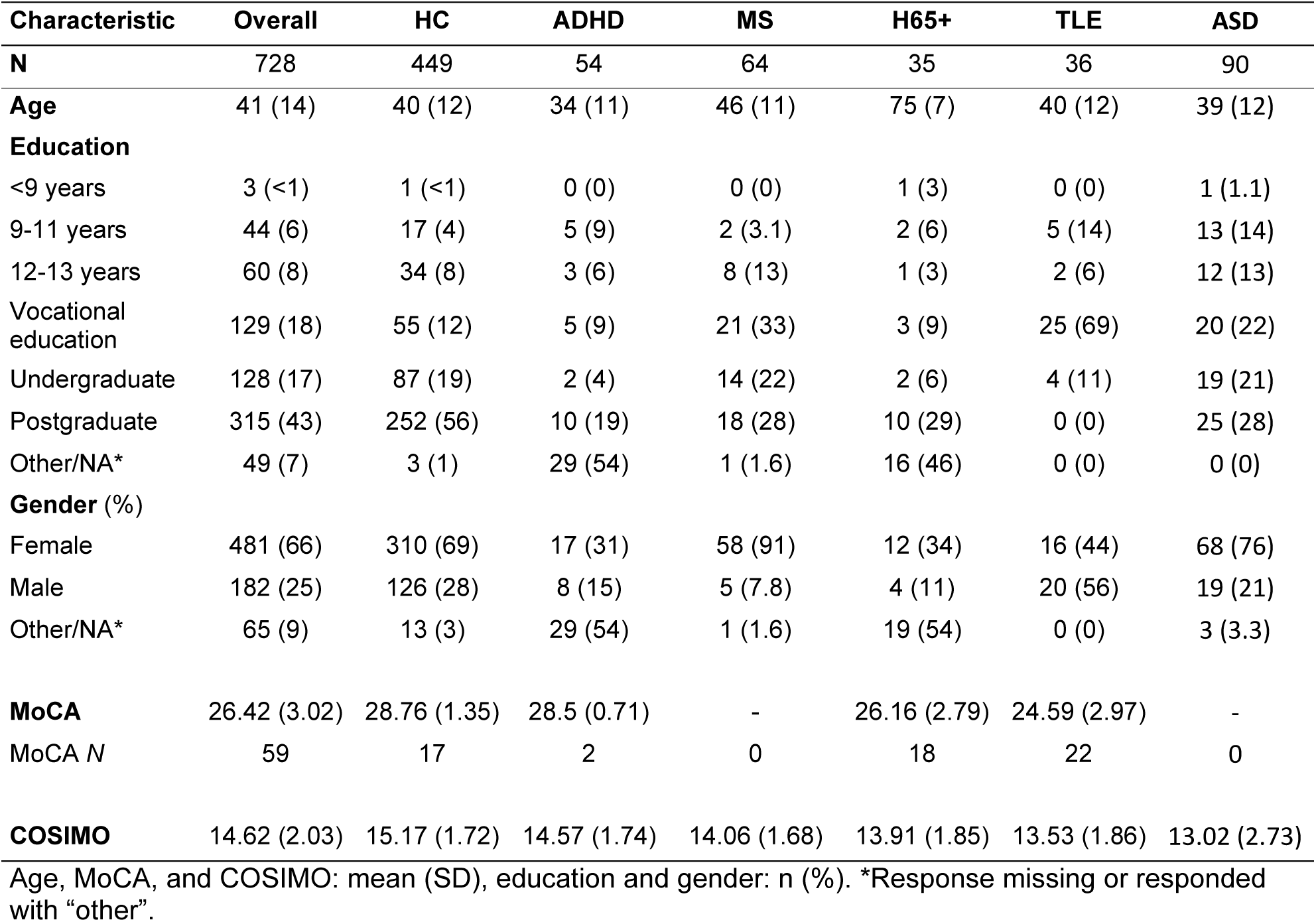
Demographic and test information.

| Characteristic | Overall | HC | ADHD | MS | H65+ | TLE | ASD |
| --- | --- | --- | --- | --- | --- | --- | --- |
| **N** | 728 | 449 | 54 | 64 | 35 | 36 | 90 |
| **Age** | 41 (14) | 40 (12) | 34 (11) | 46 (11) | 75 (7) | 40 (12) | 39 (12) |
| **Education** |  |  |  |  |  |  |  |
| <9 years | 3 (<1) | 1 (<1) | 0 (0) | 0 (0) | 1 (3) | 0 (0) | 1 (1.1) |
| 9-11 years | 44 (6) | 17 (4) | 5 (9) | 2 (3.1) | 2 (6) | 5 (14) | 13 (14) |
| 12-13 years | 60 (8) | 34 (8) | 3 (6) | 8 (13) | 1 (3) | 2 (6) | 12 (13) |
| Vocational education | 129 (18) | 55 (12) | 5 (9) | 21 (33) | 3 (9) | 25 (69) | 20 (22) |
| Undergraduate | 128 (17) | 87 (19) | 2 (4) | 14 (22) | 2 (6) | 4 (11) | 19 (21) |
| Postgraduate | 315 (43) | 252 (56) | 10 (19) | 18 (28) | 10 (29) | 0 (0) | 25 (28) |
| Other/NA* | 49 (7) | 3 (1) | 29 (54) | 1 (1.6) | 16 (46) | 0 (0) | 0 (0) |
| **Gender (%)** |  |  |  |  |  |  |  |
| Female | 481 (66) | 310 (69) | 17 (31) | 58 (91) | 12 (34) | 16 (44) | 68 (76) |
| Male | 182 (25) | 126 (28) | 8 (15) | 5 (7.8) | 4 (11) | 20 (56) | 19 (21) |
| Other/NA* | 65 (9) | 13 (3) | 29 (54) | 1 (1.6) | 19 (54) | 0 (0) | 3 (3.3) |
| **MoCA** | 26.42 (3.02) | 28.76 (1.35) | 28.5 (0.71) | - | 26.16 (2.79) | 24.59 (2.97) | - |
| MoCA *N* | 59 | 17 | 2 | 0 | 18 | 22 | 0 |
| **COSIMO** | 14.62 (2.03) | 15.17 (1.72) | 14.57 (1.74) | 14.06 (1.68) | 13.91 (1.85) | 13.53 (1.86) | 13.02 (2.73) |

Age, MoCA, and COSIMO: mean (SD), education and gender: n (%). \*Response missing or responded with "other".

The groups varied significantly in terms of age, gender, and education. Therefore, we checked whether the CER score was dependent on other factors recorded in the HC group. The effect was negligible and insignificant in the age range of 18-65 (*r* = −0.07, 95% CI −0.16 to 0.02, *p* = 0.141) using Pearson’s *r,* and for gender (*g* = 0.11, 95% CI −0.10 to 0.31, t = 0.97, *p* = 0.334) using Hedge’s *g* and a two-tailed t-test. We also found a negligible effect of education (ρ = 0.04, *p* = 0.348) using Spearman’s *ρ*. To examine the effect of general cognitive disabilities on the CER score, we performed Pearson’s correlation in the clinical groups, excluding the HC group due to a strong ceiling effect, and two extreme outliers from the clinical groups. The remaining group consisted of 17 H65+, 19 TLE, and 2 ADHD participants (*m* = 26.18, *min* = 22, *max* = 30). We found no significant correlation between the CER score and general cognitive disability (r = 0.20, 95% CI −0.12 to 0.49, *p* = 0.214).

The mean CER scores for each group are reported in Table 1. Welch’s t-test revealed that all clinical groups performed significantly worse than HC in COSIMO. Hedge’s *g* calculations for each group compared to HC revealed a small effect size for ADHD (*g* > 0.2), medium effect sizes for MS and H65+ (*g* > 0.5), and large effect sizes for TLE and ASD (*g* > 0.8), as can be derived from figure 1.

We inspected the prevalence in each group for having a score of 1.5 SD (minor impairment) or 2 SD (major impairment) below the mean of the HC group (figure 2). The highest prevalence was found in ASD for both cut-offs (37% < −1.5 SD; 22% < −2 SD). The H65+ group had a high prevalence of minor impairment (26% < −1.5 SD), but a comparatively small prevalence of major impairment (6% < −2 SD). In the TLE group, we observed 19% below −1.5 SD and 14% below −2 SD. The MS group had a prevalence of 14% < 1.5 SD and 5% < 2 SD, while ADHD exhibited the lowest prevalence with 11% < 1.5 SD and 7% < 2 SD.

**Figure 2.**
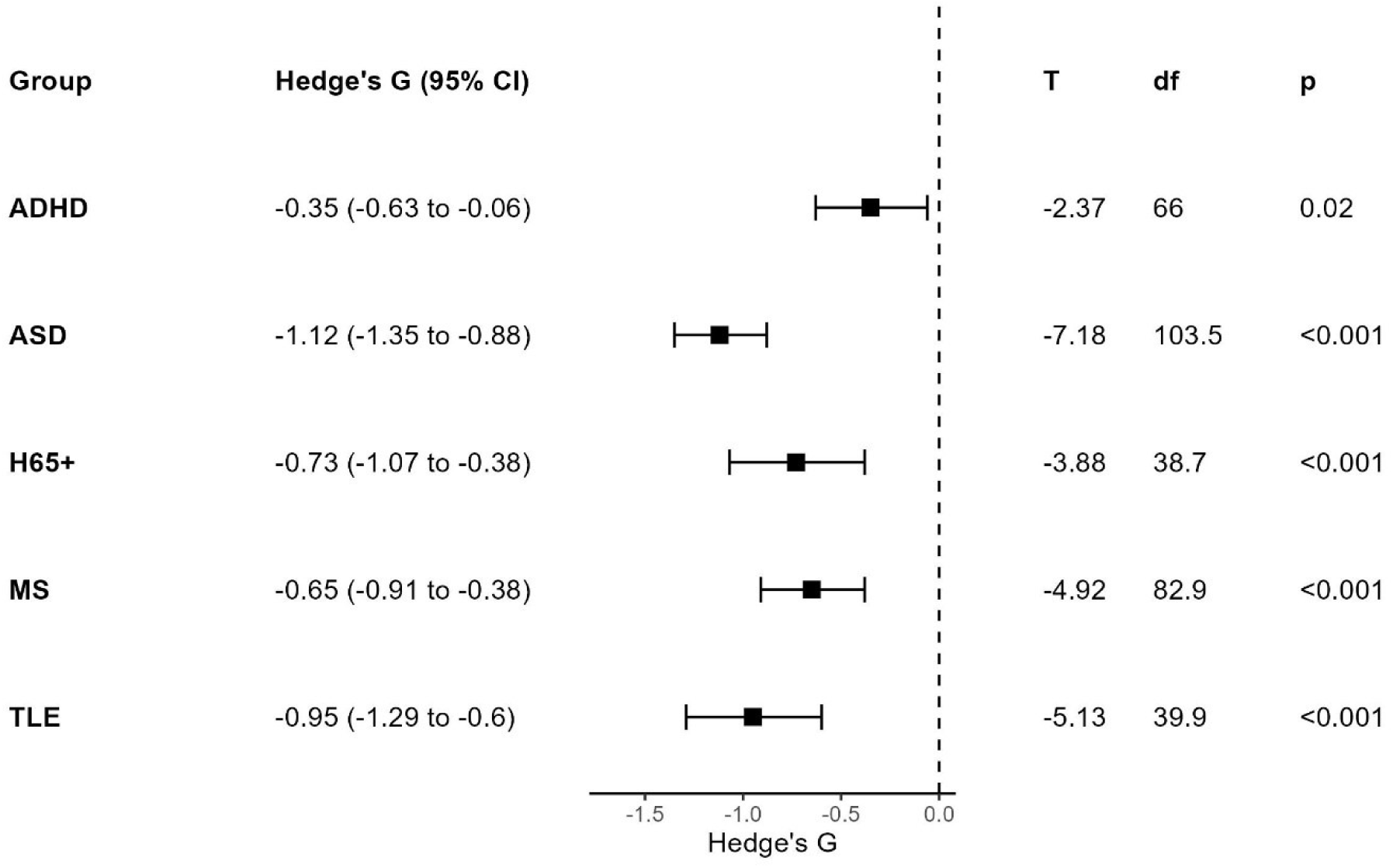
Forest plot with effect sizes and Welch’s t-test in COSIMO.

**Figure 3.**
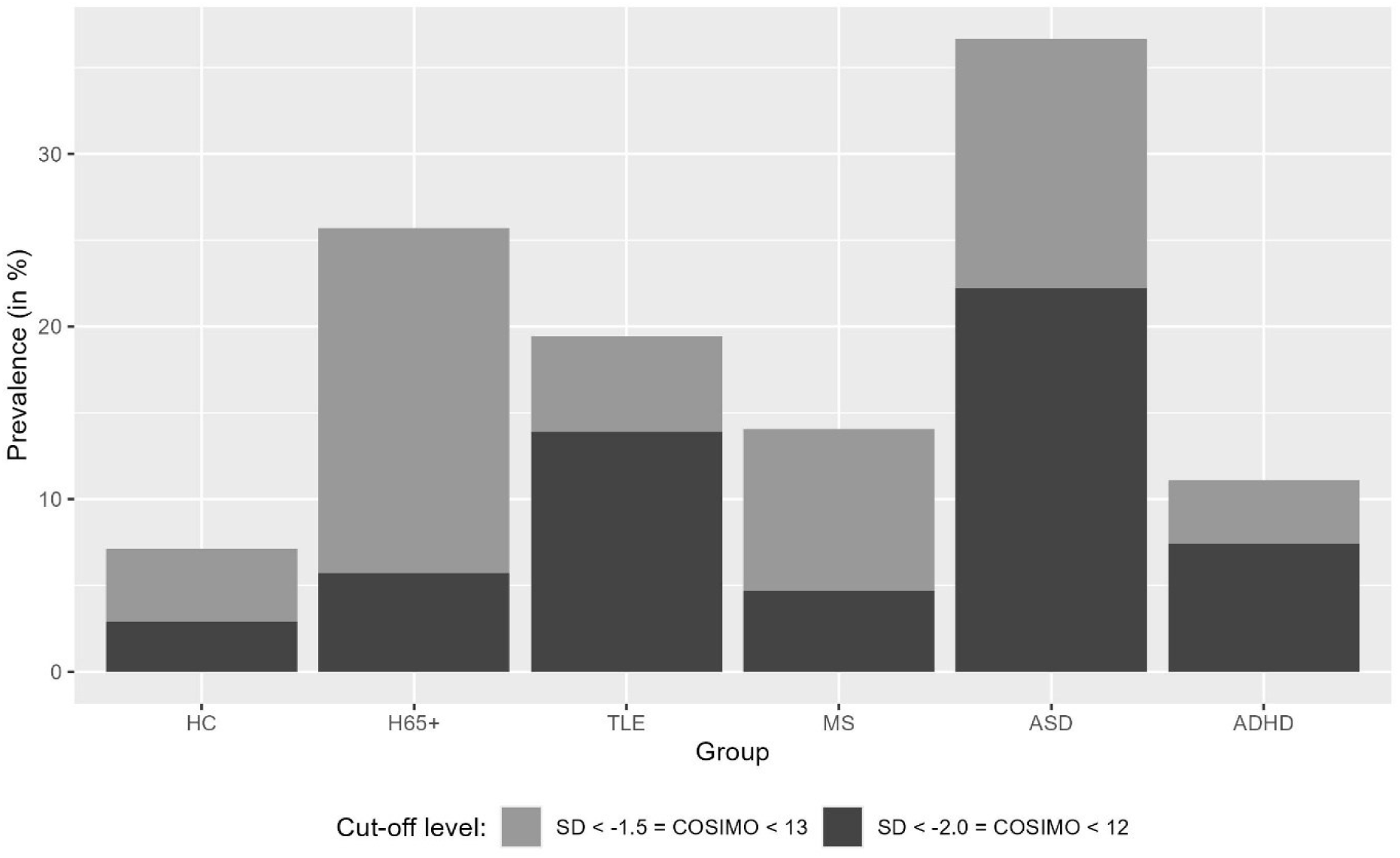
Prevalence of Complex Emotion Recognition deficits by group.

## 4 DISCUSSION

This study is the first to compare CER impairments in five clinical populations (ASD, TLE, MS, ADHD, and H65+) using a single video-based assessment method. Our findings demonstrate that CER deficits of varying severity occur in various neurological and neurodevelopmental conditions and highlight the importance of screening for social cognition in a comprehensive neurological and neuropsychiatric examination.

The level of clinical and scientific attention paid to social cognitive deficits varies greatly between the conditions involved in this study. While deficits in social functioning are part of the criteria for ASD diagnosis (Apa, 2013) and are thus well established in research and clinical practice, they are less often considered in the other conditions included in this study. Although TLE has received increased attention in social cognitive research, it has not yet been reflected in diagnostic practice (Eicher et al., 2025; Eicher and Jokeit, 2022). For MS, ADHD, and H65+, there is some evidence of social cognitive deficits, but these populations remain largely understudied in terms of social cognitive abilities, and the heterogeneous testing methods used reduce their comparability. The lack of economically feasible, ecologically valid, standardised assessment methods is a likely explanation for these gaps in our understanding of social cognitive abilities across brain pathologies (Cotter et al., 2018; Dawel et al., 2025; Eicher et al., 2025; Eicher and Jokeit, 2022).

While we see a strong effect of ASD on CER ability, consistent with the literature, the effect is weaker for ADHD. In ADHD, the attentive and affective aspect related to the prefrontal-subcortical pathology appears to impair CER abilities, whilst individuals with ASD demonstrate functional and structural anomalies in multiple brain regions important to social cognitive processing, such as the fusiform gyrus, amygdala, superior temporal sulcus (STS), and prefrontal cortex (PFC) (Liu et al., 2024). There is some evidence suggesting that autistic individuals with a comorbid ADHD have a stronger impairment in social cognition (Lievore et al., 2023). Together, these patterns indicate distinct but converging neural mechanisms, highlighting the distributed nature of social cognitive dysfunction across neurodevelopmental disorders.

The medium impairment found in H65+ adds to these findings, suggesting that pre-frontal cortical atrophy as a result of healthy aging may also cause diminished social cognitive abilities. In line with this, one meta-analysis found similar effect sizes when comparing affective theory of mind (*d* = −0.68) and emotion recognition (*d* = −0.66) in adults 50 years or older compared to younger adults (Roheger et al., 2022). Another study looking at changes in social cognition across the adult life-span found a decline in theory of mind and emotion recognition with age, independent of other cognitive functions (Grainger et al., 2023). The dependence on other cognitive functions is still debated, however, with some evidence suggesting a correlation between executive and social cognitive functioning in aging adults (Clemente et al., 2023), further underlining the pre-frontal involvement in CER.

With CER impairment found in approximately one fifth of the participants with TLE, the majority of these with scores 2SD or more below mean, our results also lend further support to the involvement of temporal and limbic structures in social cognition. The involvement of the amygdala in emotion recognition has been well established (Edmonds et al., 2024) and the superior temporal sulcus has been postulated as an important pathway for visual social perception (Pitcher and Ungerleider, 2020). Findings of a collapsed connectivity network for recognising fearful faces in TLE patients further underline the importance of these structures in social cognitive abilities (Steiger et al., 2017).

In patients with MS, it is likely that multifocal idiosyncratic abnormalities of the white matter within relevant networks are often associated with impairments in social cognition in some affected individuals. This idea is supported by our findings that approximately 14% of the tested MS population exhibited CER impairment. Only a small amount of individuals with MS exhibited major impairment (5%), indicating that, although prevalent, CER impairment is not a main feature of MS. Future studies should combine imaging and behavioural methods to better understand the neural correlates between MS and social cognitive functioning.

Our study benefited from the use of a single assessment tool with high ecological validity. We observed no significant correlation between the CER score and general cognitive functioning, indicating that the cognitive load of the CER assessment was minimal. Age, gender, and education level also appeared to have minimal to no effect on the CER score in the HC. Future studies may examine the effect of specific cognitive abilities, such as executive function compared to the CER score to further explore any potential cognitive dependencies. (Clemente et al., 2023; Lievore et al., 2023) The mixed in-clinic and online format depending on the group may have had some influence on the results, as we were unable to control for distractors in the online groups. We aimed for larger sample sizes in the groups that were mainly tested in an online setting (HC, MS, ASD) to mitigate this effect.

Further research is needed to confirm the correlation of CER with everyday functioning and quality of life, as well as to connect these findings directly to neural correlates. In addition, we need a better understanding of the effects of ageing, and how the results in healthy ageing individuals compare to pathological aging.

In conclusion, this study demonstrates that CER impairments are evident across a range of neurological and neurodevelopmental conditions, supporting the view of social cognitive impairment as a transdiagnostic feature of brain pathology. By using a single, ecologically valid paradigm, we were able to directly compare multiple clinical populations and reveal graded levels of impairment with clear distinction from HC across all groups. Our findings highlight the importance of incorporating dynamic screening of social cognition into neurological and psychiatric assessments and gives first insights into the usability of COSIMO in filling this diagnostic gap.

## CONTRIBUTORS

**Rebecca Johannessen:** Conceptualization, Investigation, Data curation, Formal analysis, Project administration, Software, Validation, Writing - original draft. **Marcel Eicher:** Investigation, Writing - Original Draft. **Anne Hansen:** Investigation, Writing - Review & Editing. **Michelle Regli:** Investigation, Writing - Review & Editing. **Lesley Ramseier:** Investigation, Writing - Review & Editing. **Tamara Hibbert:** Investigation, Writing - Review & Editing. **Hannah Sievers:** Investigation, Writing - Review & Editing. **Mara Ruepp:** Investigation, Writing - Review & Editing. **Sarah Scheibelhofer:** Investigation, Writing - Review & Editing. **Lukas Imbach:** Supervision, Writing - review & editing. **Hennric Jokeit:** Supervision, Conceptualization, Funding acquisition, Resources, Writing - Original Draft.

## ETHICS APPROVAL

This study involves human participants and was approved by the Ethics Committee of the Philosophical Faculty at the University of Zurich. Participants gave informed consent to participate in this study before taking part or consented to use of the data collected during their clinical assessment at the Swiss Epilepsy Centre.

## DATA AVAILABILITY STATEMENT

Data are available upon reasonable request. Anonymised data underlying this article will be shared on reasonable request from any qualified investigator who wants to analyse questions that are related to the published article.

## ABBREVIATIONS

ADHD: Attention Deficit Hyperactivity Disorder
ASD: Autism Spectrum Disorder
CER: Complex Emotion Recognition
COSIMO: Cognition of Social Interaction in Movies
H65+: Healthy participants aged 65 or older
MS: Multiple Sclerosis
TLE: Temporal Lobe Epilepsy

## Data Availability

https://cosimo-project.com

## REFERENCES

Apa, 2013. Diagnostic an Statistical Manual of Mental Disorders, 5th ed. American Psychiatric Association.

Baron-Cohen, S., Wheelwright, S., Hill, J., Raste, Y., Plumb, I., 2001. The ‘Reading the Mind in the Eyes’ Test revised version: a study with normal adults, and adults with Asperger syndrome or high-functioning autism. J. Child Psychol. Psychiatry 42, 241–251. 10.1111/1469-7610.00715

Bauer, J., Steiger, B.K., Kegel, L.C., Eicher, M., König, K., Baumann-Vogel, H., Jokeit, H., 2023. A comparative study of social cognition in epilepsy, brain injury, and Parkinson’s disease. Psych J. 10.1002/pchj.650

Bora, E., Pantelis, C., 2016. Meta-analysis of social cognition in attention-deficit/hyperactivity disorder (ADHD): comparison with healthy controls and autistic spectrum disorder. Psychol. Med. 46, 699–716. 10.1017/S0033291715002573

Borhani, K., Nejati, V., 2018. Emotional face recognition in individuals withattention-deficit/hyperactivity disorder: a review article. Dev. Neuropsychol. 43, 256–277. 10.1080/87565641.2018.1440295

Broicher, S.D., Kuchukhidze, G., Grunwald, T., Krämer, G., Kurthen, M., Jokeit, H., 2012. ‘Tell me how do I feel’--emotion recognition and theory of mind in symptomatic mesial temporal lobe epilepsy. Neuropsychologia 50, 118–128. 10.1016/j.neuropsychologia.2011.11.005

Chalah, M.A., Ayache, S.S., 2017. Deficits in social cognition: An unveiled signature of multiple sclerosis. J. Int. Neuropsychol. Soc. 23, 266–286. 10.1017/S1355617716001156

Clemente, L., Gasparre, D., Alfeo, F., Battista, F., Abbatantuono, C., Curci, A., Lanciano, T., Taurisano, P., 2023. Theory of Mind and Executive Functions in individuals with mild Cognitive Impairment or healthy aging. Brain Sci. 13. 10.3390/brainsci13101356

Cotter, J., Firth, J., Enzinger, C., Kontopantelis, E., Yung, A.R., Elliott, R., Drake, R.J., 2016. Social cognition in multiple sclerosis: A systematic review and meta-analysis. Neurology 87, 1727–1736. 10.1212/WNL.0000000000003236

Cotter, J., Granger, K., Backx, R., Hobbs, M., Looi, C.Y., Barnett, J.H., 2018. Social cognitive dysfunction as a clinical marker: A systematic review of meta-analyses across 30 clinical conditions. Neurosci. Biobehav. Rev. 84, 92–99. 10.1016/j.neubiorev.2017.11.014

Dawel, A., Krumhuber, E.G., Palermo, R., 2025. Faking it isn’t making it: Research needs spontaneous and naturalistic facial expressions. Affect. Sci. 10.1007/s42761-025-00320-1

Edmonds, D., Salvo, J.J., Anderson, N., Lakshman, M., Yang, Q., Kay, K., Zelano, C., Braga, R.M., 2024. The human social cognitive network contains multiple regions within the amygdala. Sci. Adv. 10, eadp0453. 10.1126/sciadv.adp0453

Eicher, M., Johannessen, R., Jokeit, H., 2025. Social neuropsychology of epilepsy in the digital age: A narrative review on challenges and opportunities. Epilepsy Behav. 166, 110336. 10.1016/j.yebeh.2025.110336

Eicher, M., Jokeit, H., 2022. Toward social neuropsychology of epilepsy: a meta-analysis on social cognition in epilepsy phenotypes and a critical narrative review on assessment methods. Acta Epileptologica 4. 10.1186/s42494-022-00093-1

Fusar-Poli, P., Placentino, A., Carletti, F., Landi, P., Allen, P., Surguladze, S., Benedetti, F., Abbamonte, M., Gasparotti, R., Barale, F., Perez, J., McGuire, P., Politi, P., 2009. Functional atlas of emotional faces processing: A voxel-based meta-analysis of 105 functional magnetic resonance imaging studies. Journal of Psychiatry and Neuroscience.

Grainger, S.A., Crawford, J.D., Riches, J.C., Kochan, N.A., Chander, R.J., Mather, K.A., Sachdev, P.S., Henry, J.D., 2023. Aging is associated with multidirectional changes in social cognition: Findings from an adult life-span sample ranging from 18 to 101 years. J. Gerontol. B Psychol. Sci. Soc. Sci. 78, 62–72. 10.1093/geronb/gbac110

Henry, J., Grainger, S., von Hippel, W., 2022. Determinants of social cognitive aging: Predicting resilience and risk. Annu. Rev. Psychol. 10.1146/annurev-psych-033020-121832

Johannessen, R., Eicher, M., Jokeit, H., n.d. Validation and norming of COSIMO: A screening tool for complex emotion recognition.

Jokeit, H., Eicher, M., Ives-Deliperi, V., 2018. Toward social neuropsychology of epilepsy: a review on social cognition in epilepsy. Acta Epilepsy 1, 8–17.

Lievore, R., Crisci, G., Mammarella, I.C., 2023. Emotion recognition in children and adolescents with ASD and ADHD: A systematic review. Rev. J. Autism Dev. Disord. 10.1007/s40489-023-00407-0

Lin, X., Zhang, X., Liu, Q., Zhao, P., Zhong, J., Pan, P., Wang, G., Yi, Z., 2021. Social cognition in multiple sclerosis and its subtypes: A meta-analysis. Mult. Scler. Relat. Disord. 52, 102973. 10.1016/j.msard.2021.102973

Liu, J., Chen, H., Wang, H., Wang, Z., 2024. Neural correlates of facial recognition deficits in autism spectrum disorder: a comprehensive review. Front. Psychiatry 15, 1464142. 10.3389/fpsyt.2024.1464142

Mangiafico, S.S., 2015. An R Companion for the Handbook of Biological Statistics, version 1.3.2 [WWW Document]. URL https://rcompanion.org/rcompanion

Marafioti, G., Cardile, D., Culicetto, L., Quartarone, A., Lo Buono, V., 2024. The impact of social cognition deficits on quality of life in multiple Sclerosis: A scoping review. Brain Sci. 14, 691. 10.3390/brainsci14070691

Nasreddine, Z.S., Phillips, N.A., Bédirian, V., Charbonneau, S., Whitehead, V., Collin, I., Cummings, J.L., Chertkow, H., 2005. The Montreal Cognitive Assessment, MoCA: a brief screening tool for mild cognitive impairment. J. Am. Geriatr. Soc. 53, 695–699. 10.1111/j.1532-5415.2005.53221.x

Pitcher, D., Ianni, G.R., Holiday, K., Ungerleider, L.G., 2023. Identifying the cortical face network with dynamic face stimuli: A large group fMRI study. bioRxiv. 10.1101/2023.09.26.559583

Pitcher, D., Ungerleider, L.G., 2020. Evidence for a Third Visual Pathway Specialized for Social Perception. Trends in Cognitive Sciences.

Pöttgen, J., Dziobek, I., Reh, S., Heesen, C., Gold, S.M., 2013. Impaired social cognition in multiple sclerosis. J. Neurol. Neurosurg. Psychiatry 84, 523–528. 10.1136/jnnp-2012-304157

Qi, L., Zhao, J., Zhao, P., Zhang, H., Zhong, J., Pan, P., Wang, G., Yi, Z., Xie, L., 2022. Theory of mind and facial emotion recognition in adults with temporal lobe epilepsy: A meta-analysis. Front. Psychiatry 13, 976439. 10.3389/fpsyt.2022.976439

Roheger, M., Brenning, J., Riemann, S., Martin, A.K., Flöel, A., Meinzer, M., 2022. Progression of socio-cognitive impairment from healthy aging to Alzheimer’s dementia: A systematic review and meta-analysis. Neurosci. Biobehav. Rev. 140, 104796. 10.1016/j.neubiorev.2022.104796

Steiger, B.K., Jokeit, H., 2017. Why epilepsy challenges social life. Seizure 44, 194–198. 10.1016/j.seizure.2016.09.008

Steiger, B.K., Muller, A.M., Spirig, E., Toller, G., Jokeit, H., 2017. Mesial temporal lobe epilepsy diminishes functional connectivity during emotion perception. Epilepsy Res. 134, 33–40.

Wang, H., Zhao, P., Zhao, J., Zhong, J., Pan, P., Wang, G., Yi, Z., 2022. Theory of mind and empathy in adults with epilepsy: A meta-analysis. Front. Psychiatry 13, 877957. 10.3389/fpsyt.2022.877957

Wang, W.-H., Shih, Y.-H., Yu, H.-Y., Yen, D.-J., Lin, Y.-Y., Kwan, S.-Y., Chen, C., Hua, M.-S., 2015. Theory of mind and social functioning in patients with temporal lobe epilepsy. Epilepsia 56, 1117–1123. 10.1111/epi.13023

Wolf, I., Dziobek, I., Heekeren, H.R., 2010. Neural correlates of social cognition in naturalistic settings: A model-free analysis approach. Neuroimage 49, 894–904. 10.1016/j.neuroimage.2009.08.060

Yeung, M.K., 2022. A systematic review and meta-analysis of facial emotion recognition in autism spectrum disorder: The specificity of deficits and the role of task characteristics. Neurosci. Biobehav. Rev. 133, 104518. 10.1016/j.neubiorev.2021.104518

Ziaei, M., Arnold, C., Thompson, K., Reutens, D.C., 2023. Social cognition in temporal and frontal lobe epilepsy: Systematic review, meta-analysis, and clinical recommendations. J. Int. Neuropsychol. Soc. 29, 205–229. 10.1017/S1355617722000066

